# Retinal fundus photographs capture hemoglobin loss after blood donation

**DOI:** 10.1101/2021.12.30.21268488

**Authors:** Akinori Mitani, Ilana Traynis, Preeti Singh, Greg S. Corrado, Dale R. Webster, Lily H. Peng, Avinash V. Varadarajan, Yun Liu, Naama Hammel

**Affiliations:** Google Health, Palo Alto, CA, USA; Work done at Google Health via Advanced Clinical, Deerfield, IL, USA

## Abstract

Recently it was shown that blood hemoglobin concentration could be predicted from retinal fundus photographs by deep learning models. However, it is unclear whether the models were quantifying current blood hemoglobin level, or estimating based on subjects’ pretest probability of having anemia. Here, we conducted an observational study with 14 volunteers who donated blood at an on site blood drive held by the local blood center (ie, at which time approximately 10% of their blood was removed). When the deep learning model was applied to retinal fundus photographs taken before and after blood donation, it detected a decrease in blood hemoglobin concentration within each subject at 2-3 days after donation, suggesting that the model was quantifying subacute hemoglobin changes instead of predicting subjects’ risk. Additional randomized or controlled studies can further validate this finding.

## Introduction

Machine learning has been increasingly applied to medical images for novel tasks such as predicting cardiovascular risk factors, coronary artery calcium scores, and detecting chronic kidney disease and type 2 diabetes from retinal fundus photographs.^1–4^ In another study,^5^ deep learning models were able to predict blood hemoglobin concentration (ie, Hb concentration, or “[Hb]”) from retinal fundus photographs at an accuracy (mean absolute error (MAE) of 0.67 g/dL) comparable to invasive testing by a point-of-care hemoglobin analyzer, suggesting potential utility as a cost-effective add-on to established diabetic retinopathy screening programs. In this study, the model was validated using data from the UK Biobank, where fundus photographs and associated lab measurements of [Hb] were available. Independent investigators further verified that the model generalized; ie, it was able to predict [Hb] and anemia in patients from a different continent and using different imaging devices.^6^

However, these studies were limited to images collected at only one time point per patient. As such, the validation design of the prior studies did not enable evaluation focused on [Hb] change. Therefore, the models could be predicting [Hb] using “constant” or non-modifiable characteristics of the subjects, such as age, self-reported sex, or ethnicity (all of which are correlated with [Hb]^7,8^ and can be predicted to some extent from fundus photographs^1,9^) as discussed previously^10^. While the previous study showed that the model predicted [Hb] more accurately than using demographic information, the performance improvement may still be attributed to other non-modifiable factors such as anatomic phenotypes. Better understanding of whether the models are measuring current [Hb] (as opposed to a statistical “prior” on [Hb] based on patients’ pretest probability of anemia or average [Hb] given demographic variables) is crucial to understanding its limitations and appropriate real world use. For example, models that measure anemia likelihood would be unlikely to help track anemia progression or recovery via repeated retinal photographs because the predictions would remain relatively constant.

To distinguish between these possibilities, we conducted a non-randomized, prospective, pre-post observational study to investigate whether the model-predicted [Hb] is affected by an intervention that acutely lowers [Hb]. Specifically, we recruited subjects who were volunteering for blood donation, and used the deep learning model to examine the retinal fundus photographs taken before versus after blood donation. Blood donation itself does not change the [Hb] (ie, the *concentration*) immediately, because the decrease in total Hb mass is balanced by a corresponding decrease in the circulating blood volume.^11,12^ The natural progression of physiological homeostasis following blood loss causes a delayed hemodilution upon blood plasma replenishment, which causes a [Hb] “#$%#&’# ()&(*#&+’ &(& “%,*, - ./ 01”2 &-(#% ‘#3#%&4 “&5’6^11,12^ The subsequent [Hb] recovery requires red blood cell regeneration and takes a few months to a half year.^12^ Thus, blood donation poses a unique opportunity to examine the effect of subacute [Hb] decrease.

## Results

In this study, we captured fundus photographs at 3 time points: (1) within an hour before blood donation; (2) within an hour after blood donation; and (3) 2-3 days after blood donation (Figure 1, Methods). Four fundus photographs were taken for each time point (left and right eyes, macular-centered and primary fields), and the [Hb] predictions for the four images were averaged per participant for comparison across time points using the Wilcoxon signed-rank test and with Bonferroni correction (Figure 2). The difference between predicted [Hb] before blood donation and after blood donation on the same day was not statistically significant (Figure 2b; p = 0.056; absolute change: -0.25 ± 0.08 g/dL [mean ± standard error], n=14). The predicted [Hb] 2-3 days after the blood donation was also not statistically significantly different compared to after blood donation on the same day (absolute change: -0.26 ± 0.09 g/dL; Figure 2c, p = 0.091), but was statistically significantly different compared to before blood donation (absolute change: -0.51 ± 0.10 g/dL; Figure 2d, p = 0.009). The predicted [Hb] at the first time point was compared against the reported [Hb] before the blood donation, and the MAE was 0.76 (95% confidence interval: 0.46 to 1.06, n=10). This is similar to previously reported MAE of 0.67 in the UK Biobank population.^5^

**Figure 1.**
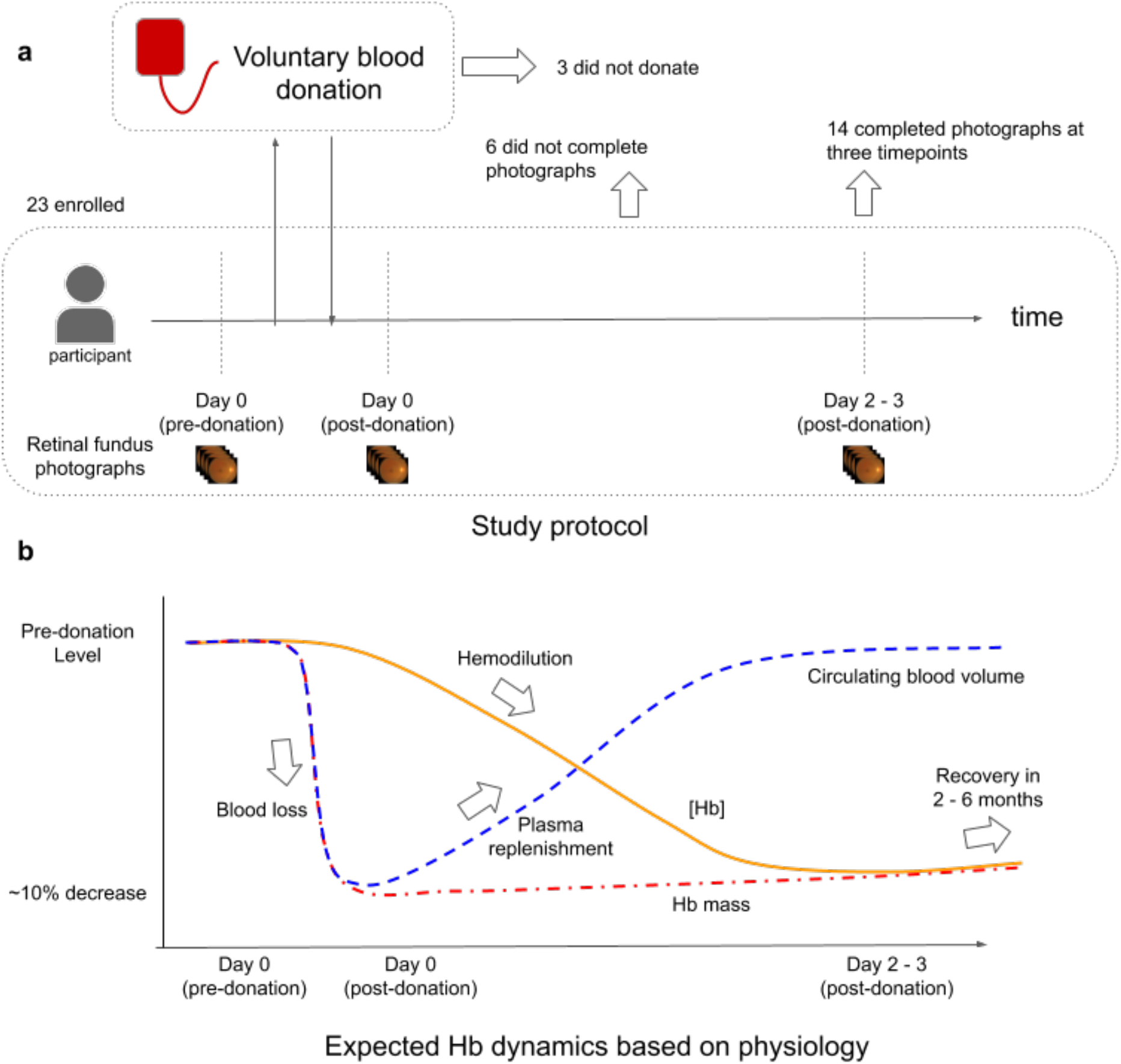
**a**, Study design schematic, showing enrollment of volunteers who were scheduled to donate blood at a blood drive. Fundus photographs were taken at three time points: before the donation, within 1-2 hours after the donation, and 2-3 days post-donation. Participants who did not end up donating blood, or did not complete the fundus photography sequence were excluded. **b**, Based on physiology,^11–13^ the expected change in blood volume (blue broken line), hemoglobin (Hb) mass (red dash-dotted line), and hemoglobin concentration ([Hb], orange solid line) change over time post-donation. While total Hb mass decreases immediately, the [Hb] does not decrease until the lost plasma is replenished over the course of a few days. The recovery of [Hb] requires actual red cell regeneration and is a gradual process that takes months.

**Figure 2.**
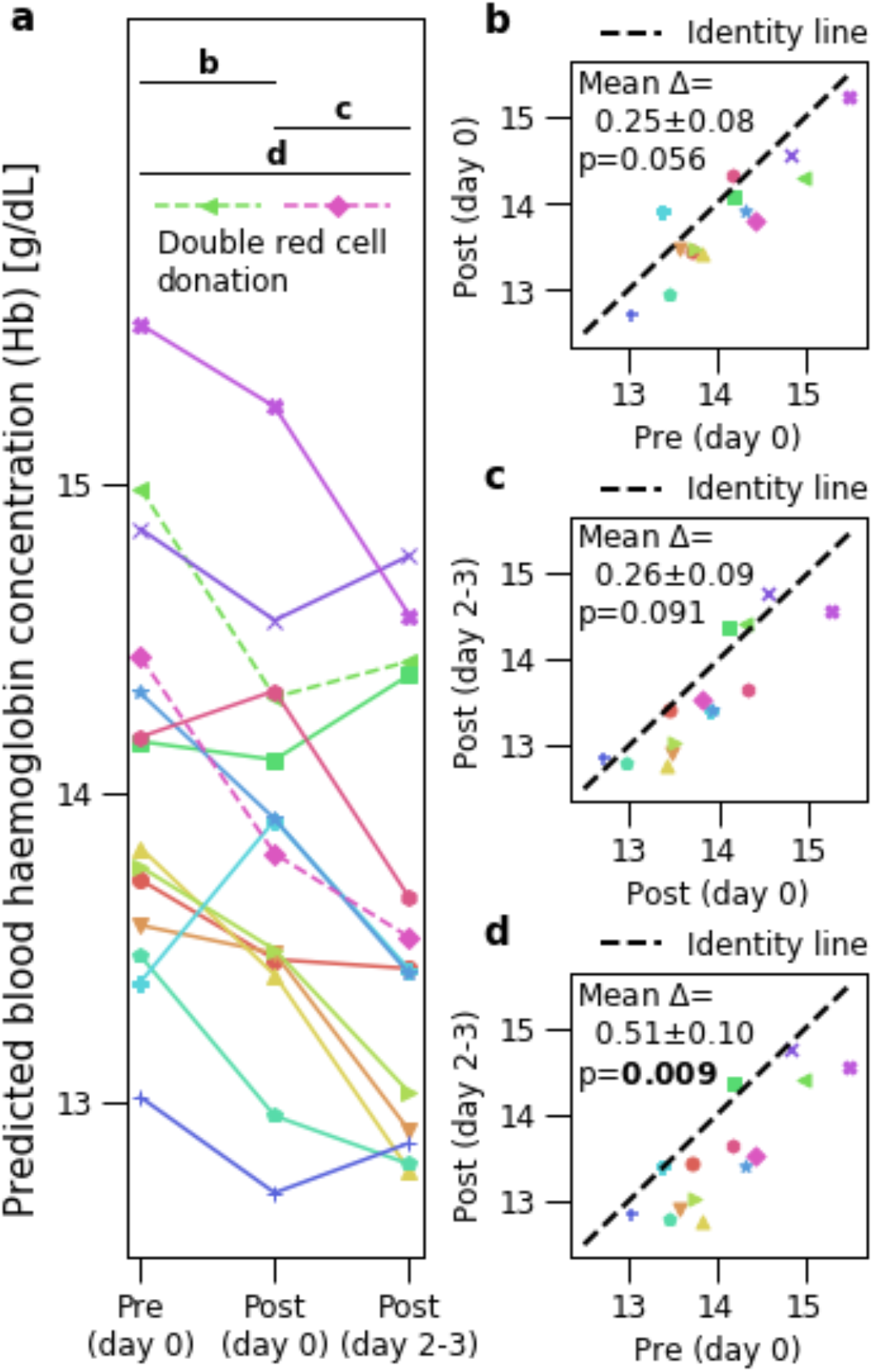
**a**, Mean predicted blood hemoglobin concentration ([Hb]) for each participant at different timepoints. Broken lines indicate participants who reported double red cell donation. The three horizontal black lines indicate the three pairwise comparisons shown in panels **b**-**d. b**, Difference in predicted Hb between before and after blood donation on the same day. A black broken line indicates the identity line, where the predictions are the same in the two conditions. Text shows p-value (Wilcoxon signed rank test with Bonferroni correction), and mean and standard error of differences (pre-post, n=14). **c**, same as **b**, but comparing after blood donation on the same day and 2-3 days after blood donation. **d**, same as **b**, but comparing before blood donation and 2-3 days after blood donation. Bold indicates a statistically significant p-value.

## Discussion

The results showed a decrease in model-predicted blood [Hb] after blood donation. For a standard whole blood donation, a participant with 5000 mL of total blood would lose approximately 10% of their total hemoglobin in a 475 mL (1 pint) blood donation. This would lead to an approximately 1 g/dL decrease in [Hb] over a few days, once the blood plasma volume equilibrates (Figure 1).^11–13^ In this study, [Hb] decrease was estimated theoretically instead of measured invasively so as to avoid an unnecessary additional invasive procedure (venous blood draw or finger prick). The observed mean difference of the predictions (0.51 ± 0.10 g/dL) was comparable to the theoretical calculations. The slight difference can potentially be attributed to the proportional bias of the model;^5^ the model overestimates low [Hb], which may result in an underestimation of the magnitude of the decrease. Improved calibration may help in this regard, and indeed the previously-described recalibrated model^5^ increases the mean difference of predicted [Hb] to 0.74 ± 0.15 g/dL.

Interestingly, the model predictions also decreased (albeit by a much smaller magnitude on average, and not meeting statistical significance) for images taken on the same day after blood donation. In theory, the immediate effect of blood donation is on blood volume instead of concentration, suggesting that the model’s predictions may also be affected by total blood volume. Further validation on a larger cohort would be necessary to validate this finding, to evaluate appropriateness of application of this technique to situations where subjects may have experienced acute blood loss.

When machine learning models are developed and evaluated using data from a single time point per subject, potential mechanisms underlying a significant association include not only direct measurement of the current condition, but also features that reflect pre-existing conditions or indicate risk of developing the current condition. For example, increased retinal venous tortuosity is associated with a longer duration of anemia.^14^ If a model only uses features that take some time to develop, the prediction indicates that the condition has been present for a while (and could have already resolved) instead of the current state. Hypothetically, if the subjects need to be anemic for months before the features used by the model manifests, the model should not immediately capture a [Hb] decrease following blood loss from blood donation. Though the detection of delayed responses can be a desirable feature in some applications (similar to how HbA1c is used for tracking average diabetes control over a few months), our results suggest detection of delayed responses is unlikely.

Alternatively, the model may be detecting features related to “constant” or unmodifiable risk factors (e.g. genetic information, age, or anatomical phenotype). Such a model may be useful for screening, but repeated measurements cannot be used to track disease progression or treatment response. Therefore, differentiating between these possibilities and examining the temporal response of the model predictions is critical in assessing the clinical value of the machine learning models.

This pre-post study design was designed to avoid additional invasive procedures (eg, venous blood draws or finger pricks to quantify [Hb]) and intervening in the blood donation process (eg, via a randomized control arm where control subjects would be instructed not to donate). A similar pre-post study using transfusions (ie, adding instead of removing blood) was also conducted as a subanalysis in a prior work leveraging deep learning to predict the presence of anemia from electrocardiograms.^15^ However, the constraints of this study design means that some of the trends observed could be due to other unforeseen differences before and after the intervention (ie, between day 0 and days 2-3). Future studies may leverage the data from this pilot to help justify the utilization of venous draws or a reduction in the total number of blood donors in a randomized controlled study. Another potential design is a case-control study where subjects who donated blood could be matched with controls based on age and sex. In such a case-control study, the control arm would help determine whether other confounding factors contribute to the observed trends. As such, future studies are needed to confirm our observations.

To summarize, this study utilized voluntary blood donation as an intervention that induces [Hb] decrease in participants, and confirmed that the model could capture short-term, time-varying changes in [Hb]. This observation suggests a low likelihood of a long temporal delay in model detection of [Hb] changes, or that the model is solely predicting [Hb] by quantifying pretest likelihood of anemia. Still, there remain open questions about the temporal response of the model, such as if the model would over- or underestimate [Hb] in the setting of an ongoing anemia progression. Further study is needed to evaluate the model’s ability to track [Hb] changes or anemia progression and treatment response.

## Methods

### Participants and study design

Adult participants (age >= 18 years) planning to donate blood to a blood bank were recruited to volunteer for this study. The study was reviewed and approved by Advarra Institutional Review Board. All participants signed an approved consent form prior to initiating study activities and no compensation was offered for their participation.

Each participant provided consent, completed a questionnaire, and underwent retinal fundus photography at three time points. Fundus photographs were taken at all three time points in a dark setting without pupil dilation using a Topcon NW-400. Age range and sex were self-reported at study enrollment (the first time point). After 1-3 hours (the second time point), the participants were asked if they had donated blood (whole blood donation of approximately 475mL, double red cell donation, plasma-only donation, or no donation). Participants also reported their pre-donation point-of-care blood [Hb], which was measured by the blood bank prior to blood donation. At the first two time points, the fundus photographs were taken in a trailer located near the blood bank vehicle. After 3 days (the third time point), the fundus photographs were taken in a darkroom.

Participants who reported no blood donation or plasma donation only were excluded from the analysis (n=3). Participants who reported double red cell donation were included in the analysis (n=2). To qualify for whole blood donation, donors must satisfy requirements such as feeling well, weighing at least 50 kg, and having [Hb] of at least 12.5 g/dL (female) or 13.0 g/dL (male).^16^ Participants who were unable to complete the full sequence of retinal fundus photographs at the three time points were excluded from the analysis (n=6). Because of scheduling reasons, 1 participant had the third time point’s images taken after 2 days (instead of 3 days). After the exclusions, 14 participants were included in the analysis. Of these, the self-reported age ranges were: 7 between 20-29, 4 between 30-39, and 3 above 39. The self-reported sex were: 9 male and 5 female. There were no observable image quality differences between the 3 time points.

### Model development

The model used in this study was previously described.^5^ Briefly, an ensemble of artificial neural networks based on the Inception-v3 architecture were trained to predict variables from the complete blood count (red blood cell count, hematocrit, and hemoglobin concentration) using data from the UK Biobank.^17^ Only the hemoglobin concentration predictions were used in this study.

### Statistical analysis

To test for differences across different time points, the Wilcoxon signed-rank test was used. Bonferroni correction was applied to adjust the p-values for the three comparisons conducted in this analysis. To obtain 95% confidence intervals for MAE, we used the bootstrap procedure with 2,000 samples and reported the 2.5 and 97.5 percentiles.

## Data Availability

The informed consent and study protocol does not permit study images to be made available.

## Data availability

The informed consent and study protocol does not permit study images to be made available. Researchers interested in validating this model on their data can contact the corresponding author regarding a research collaboration (N.H.)

## Acknowledgements

We would like to acknowledge Dr. Suchitra Pandey, Clayton Toller, and Renee Gibson at the Stanford Blood Center for their guidance and help in planning the study around a scheduled blood drive. Gratitude also goes to Tayyeba Ali, Siva Balasubramanian, Himanshu Chavda, Benny Ayalew, Joy Lavigne, Noemi Figueroa, Saumya Pandey, Leslie Brody, and Alejandra Maciel for their assistance in planning or conducting the study.

## Author contributions

A.M., I.T., and N.H., designed the study. A.M., I.T., P.S., N.H., executed the study. A.M., I.T., Y.L. and A.V.V. analysed the data and interpreted the results. A.M., I.T., Y.L. and N.H. drafted the manuscript with inputs from all authors. G.S.C., D.R.W., L.P., and A.V.V provided strategic guidance and oversight. All authors contributed to manuscript revision and approved the submitted version.

